# Autoimmune encephalitis patients identified among individuals attending Swedish memory clinics

**DOI:** 10.64898/2026.02.09.26345301

**Authors:** A Freitas-Huhtamäki, N Kleebauer, A Gardner, J Lundberg, M Esbjörnsson, R Da Silva Rodrigues, Patrick Waters, M Scheller-Nissen, M Blaabjerg, N Bogdanovic, J Theorell

**Author notes:** **Correspondence to:**, Center for Infectious Medicine, Department of Medicine Huddinge, Karolinska Institutet, Stockholm, Sweden; Mailing address: CIM/ANA Futura, Alfred Nobles Allé 8B, 141 52 Huddinge, Karolinska Institutet.

## Abstract

**Introduction:** Neurodegenerative dementia syndromes are severely debilitating, progressive and increasing in incidence with an ageing population. A treatable differential diagnosis to neurodegenerative dementia is autoimmune encephalitis (AE), but AE patients are often misdiagnosed, delaying treatment. Previous work in the Netherlands has shown that 0.8% of patients with suspected neurodegenerative dementia suffer from AE. In Sweden, there is considerable variability in the prevalence of AE, possibly indicating under-diagnosis. We hypothesized that some Swedish individuals seeking care for memory impairment might suffer from an undetected AE and that these would show aberrances in available markers of neuroinflammation.

**Methods:** We retrospectively screened frozen sera from 1041 individuals seen between 2019 and 2023 at the Karolinska University hospital memory clinics in Stockholm for autoantibodies to contactin-associated protein-like 2 (CASPR2), leucine-rich glioma-inactivated 1 (LGI1), gamma-aminobutyric acid receptor B (GABA_B_R), the n-methyl-d-aspartate receptor (NMDA-R) and Immunoglobulin superfamily containing LAMP, OBCAM, and Neurotrimin family member 5 (IgLON5) using live cell-based assays (CBAs) and scored them by microscopy. Serum and CSF from suspected positive patient samples were re-tested and titrated by live CBA, commercial fixed CBAs and tissue based assays.

**Results:** 8 of the 1021 individuals, or 0.8% of the cohort, tested positive in at least three different tests for antibodies to CASPR2 (n=3), GABABR (n=2), LGI1 (n=1) and NMDAR (n=2). Seven of these patients had not been previously diagnosed with AE. Apart from two CASPR2-antibody positive patients showing neuropathic pain and seizures and neuromyotonia, respectively, the patients lacked clinical signs of encephalitis aside from memory impairment and affect lability. The antibody-positive patients did not differ significantly from autoantibody-negative patients in any available clinical parameter. None showed signs of inflammation on brain magnetic resonance tomography, and 2/7 lacked any sign of neuroinflammation in the CSF with available tests, which is commonly seen in later-onset AE.

**Conclusion:** Our work identifies undiagnosed AE patients with subtle symptomatology among Swedish memory clinic visitors, that cannot be sensitively separated from antibody-negative patients with current diagnostic tests. Our results suggest the need for the introduction of more sensitive markers of neuroinflammation to the memory clinic to identify and treat individuals with AE among sufferers of memory impairment.

## 1. Introduction

Autoimmune encephalitides (AEs) are a group of inflammatory brain disorders where commonly IgG autoantibodies directed against extracellular epitopes on neuronal proteins alter neuronal function (1). First defined around 20 years ago (2,3), this group is expanding both in clinical presentations and in prevalence with increasing recognition (4,5). Depending on which neuronal protein that is targeted, the patients can get a wide spectrum of symptoms, such as seizures, affective lability, psychosis, movement disorders and autonomic dysregulation (1). An additional feature of AE is short-term memory impairment, which tends to increase as the disorder progresses. Complicating the matter, AEs presenting later in life tend to be more subtle and slowly progressing, increasing the risk of under-diagnosis and therefore often leading to significant memory impairment (1). In line with this, Bastiaansen *et al* showed in a cohort of 175 autoimmune encephalitis patients presenting after 45 years of age, 38% fulfilled dementia criteria without prominent seizures and 18% were initially suspected to suffer from neurodegenerative dementia (6), with the most prevalent autoantigens being contactin-associated protein-like 2 (CASPR2), leucine-rich glioma-inactivated 1 (LGI1), gamma-aminobutyric acid receptor B (GABA_B_R), and the n-methyl-d-aspartate receptor (NMDA-R) subunit 1 (GRIN1), the autoantibody target in NMDA-R encephalitis. Furthermore, the same group showed that 7/920 (0.8%) individuals with suspected neurodegenerative dementia had antibodies to neuronal surface antigens and did upon close inspection show atypical signs such as subacute onset or fluctuating symptoms, but generally lacked overt inflammation or MRI abnormalities (7). The autoantigens identified in this study were Immunoglobulin superfamily containing LAMP, OBCAM, and Neurotrimin family member 5 (IgLON5), LGI1, NMDAR-GRIN1 and Dipeptidyl-Peptidase-Like Protein-6 (DPPX).

Similar to what has been reported elsewhere (4), Sweden seems to have a geographically uneven distribution of AE cases. In a study covering a fifth of the population (∼2 million people) over five years, Kosek and coauthors estimate an annual incidence of 3.3 per million inhabitants per year, with a dominance of AE with NMDA-R and 65 kDa glutamic acid decarboxylase (GAD65) autoantibodies (8). In contrast, 5-year clinical observations in Hässleholm hospital, Skåne, Sweden, a smaller hospital with considerable AE competence, suggest higher annual incidence rates of up to 40 per million inhabitants per year in the catchment area, or 12 times higher than in the published prevalence study (clinical observations, M.E.). The diagnoses in this group is dominated by LGI1, CASPR2 and paraneoplastic central nervous system disorders, i.e. late-onset diagnoses (clinical observations, M.E.). This is thought to be due to a uneven recognition of AEs, a notion underscored by a considerable increase in the AE incidence over the last decennium, with a relatively more pronounced increase in LGI1-antibody encephalitis, which presents more subtly and later in life compared to NMDA-R encephalitis (clinical observation and (4)).

We hypothesise that, as in the Netherlands (7), some undiagnosed Swedish AE patients seek medical care because of memory-impairment. Our primary aim is therefore to define the prevalence of AE in a Swedish memory clinic cohort. Our secondary aim is to establish whether available biomarkers of neuroinflammation and neuronal damage, including CSF cell count (9), albumin quotient, kappa-free light-chain intrathecal fraction (10), CSF neurofilament light chain or CSF tau (11,12) differ between identified AE patients and other patients in the cohort.

## 2. Methodology

### 2.1. Patients and ethical approval

This study was approved by the Swedish Ethical Review Authority, registration number 2011/1987-31 and 2022/03556-01. All patients provided written consent after receiving oral and written information about the study. 1021 patients undergoing dementia diagnostics due to rapid or early-onset memory impairment between 2019 and 2023 were included. These patients were retrospectively included from GEDOC, a long-running cohort of patients coming to the memory outpatient clinics at the Karolinska University Hospital, Stockholm, Sweden(13). All patients coming to the clinic were asked to participate in the cohort and approximately 91% of the patients undergoing a lumbar puncture agreed to participation during the inclusion period. The resources available for the cohort includes a biobank of frozen serum and CSF, and a database with information about gender, age and diagnosis as well as scores from neuropsychological testing (e.g., MMSE, MoCA, RAVLT), brain MRI, APOE genotyping, and clinical biomarker analyses in CSF and blood (e.g., amyloid β42, amyloid β42/40 quotient, phospho-tau181, total tau and neurofilament). The sub-cohort used in this study was enriched for individuals with available CSF and for whom the kappa-free light chain-intrathecal fraction (KFLC-IF) had been measured. 927/1021, or 91% of the cohort had both CSF and KFLC-IF. The selected subgroup made up 55% of all patients undergoing a lumbar puncture during the time period, including those who declined participation in the cohort study.

### 2.2. Overarching design of screening and confirmation

All sera were screened for autoantibodies to CASPR2, LGI1, GABA_B_R, IgLON5 and the (NMDAR) subunit GRIN1 using in-house live cell-based assays, described in detail below. All assays were repeated at least twice for each serum. In cases where a signal above background was detected consistently in the two replicates, CSF was retrieved and both serum and CSF were sent for external confirmatory testing by commercial cell-based assays at the Karolinska University Hospital Clinical Immunology Laboratory Unit, Stockholm, Sweden, as well as in-house tissue-based assays performed by the Odense Autoimmune Neurology Group, University of Southern Denmark, Odense, Denmark.

### 2.3. Plasmids

Plasmids for CASPR2, GABABR and IgLON5 were designed with and acquired from VectorBuilder (Chicago, IL, USA) (Supplementary table 1). LGI1-CASPR2 tail fusion protein and NMDAR (GRIN1) plasmids were kindly provided by the Oxford Autoimmune Neurology Diagnostic Laboratory, University of Oxford, UK, (14,15). For CASPR2, IgLON5 and LGI1-CASPR2, eGFP was fused to the intracellular c-terminus. For the GABABR, plasmids with GABABR1 and GABABR2 constructs were used in a 5:2 proportion. The GABAB-R1 and -R2 proteins were fused to intracellular, c-terminal FLAG tags, that were not used in the project. For the NMDAR antibody detection, a plasmid containing the native GRIN1 construct was used without any GRIN2 construct. The plasmids were either bought ready to use from VectorBuilder (CASPR2, and IgLON5) or expanded and extracted from bacteria using EndoFree MaxiPrep kits (Qiagen, Venlo, the Netherlands). The competent bacteria transformations were performed by VectorBuilder for GABAB and in-house for GRIN1 and LGI1 constructs.

### 2.4. Live cell-based assay (CBA)

A detailed CBA protocol with modifications for each antigen can be downloaded at https://github.com/jtheorell/Laboratory-protocols. It is based on published protocols (Irani 2010) with a few modifications. Instead of conventional HEK-293 cells, a HEK-293 derived cell line expressing the macrophage scavenger receptor, called 293-MSR cells were used, as these more strongly adhere to plastic, and therefore are more adherent. Lipofectamine (Invitrogen) was used as a transfection agent. For GRIN1, the NMDA-R inhibitor MK801 was added after eight hours with a final concentration of 30 μM. Time from transfection to screening was 48 hours for all antigens apart from GRIN1, where 24 hours was used.

### 2.5. Screening procedure

For screening, sera were diluted 1:100 for CASPR2 and 1:20 for the remaining antigens. Negative and positive controls from the Autoimmune Neurology Diagnostic Laboratory, University of Oxford, UK (all but IgLON5) and the Autoimmune Neurology Group, University of Southern Denmark, Odense, Denmark (IgLON5) were included in all plates. To ensure a clear positive signal, the positive controls were used at concentrations at least 4 times higher than the end-point dilution. All screening was performed in duplicates. CSF was screened undiluted. For titrations of positive samples, the end-point dilution was defined as the highest dilution giving a positive score. For all antigens apart from GRIN1, the secondary antibody was a goat anti-human cross-absorbed IgG (H+L) conjugated to Alexa fluor 568 (Invitrogen). For NMDA-R, the secondary antibody was a goat anti-human Fc cross-absorbed IgG (Invitrogen), followed by a tertiary donkey-anti-goat Fc cross-adsorbed IgG conjugated to Alexa fluor 568 (Invitrogen). Finally, all wells were coated with a 4′,6-diamidino-2-phenylindole (DAPI)-containing mounting medium (ThermoFisher), staining nuclei blue.

Microscopy imaging of live cell-based assays was performed using a Nikon Ti2 inverted widefield/spinning disk CREST v3 microscope. This system was equipped with a fully motorized stage, a 10x (NA 0.45), or 20x air objective (NA 0.75), and a Kinetix sCMOS camera (>[95% quantum efficiency, 6.5 μm pixel size, 2720[×[2720-pixel field of view), including 1. Images from three random positions selected by the microscope software were captured per well, focusing on a pre-selected central area to ensure cell capture even in wells with lower confluency. The files were saved in .nd2 format, with a 16-bit intensity range with identical intensities for all images within each plate. The imaging was fully automated, including finding the focal plane. In cases where the random positions still gave a low per-well cell count, the imaging was re-done for the whole plate.

In cases where a whole plate showed low confluency, another replicate was performed until two replicates with sufficient quality were available. This created in up to four replicates for IgLON5, and up to three replicates for remaining antigens.

Images taken with this robotised microscopy approach were scored by visual inspection.

### 2.6. Experimental setup and analysis of confirmatory tissue-based assays

For tissue-based screening, adult Sprague-Dawley rats were euthanized and brains removed under aseptic conditions. Brains were immediately submersed in cold 4% paraformaldehyde in Phosphate-buffered saline (PBS), for 1 hour. After this, brains were transferred to 40% sucrose in PBS for 48 hours at 4 degrees, for cryoprotection. Brains were separated into hemispheres and individual hemispheres were encapsulated in cryoprotection media (Cryoembed – Leica microsystems) and snapfrozen in ice-cold isopentane. Individual capsules were kept at -80 degrees until sectioning. Brains were cut into 6 micrometer tissue sections and placed on microscope slides (Superfrost plus, Thermo-Fischer). Tissue sections were kept at -20 degrees until used. For immunohistochemistry, sections were thawed to room temperature (RT) and washed in TBS, followed by incubation with H_2_O_2_ in TBS for 30 minutes, then washed in TBS and blocked with blocking buffer containing 2% Bovine Serum Albumin and 5% donkey serum in TBS for 30 min. After additional washing, slides were incubated with serum (1:50) or CSF (1:2) in blocking buffer overnight at 4 degrees. The next day, slides where washed in TBS and incubated with secondary antibody (donkey anti-human IgG, 1:2000, Thermo-Fisher) for 1h at RT. Slides were developed using a standard diaminobezadine (DAB) protocol and coverslipped in DPX mounting media (Thermo-Fisher). Slides were visualized using BF microscopy and scored visually. As positive controls, we used slides incubated with purified NMDA, IgLON5 and LGI1 IgG (all 1:50) from patients with definite AE and a classic clinical phenotype.

### 2.7. Experimental setup and analysis of confirmatory, commercial cell-based assays

Confirmatory analysis of neuronal surface antibodies was performed using commercial fixed cell-based assays (Euroimmun, Lübeck, Germany) in accordance with the manufacturer’s instructions. CSF samples were tested at 1:1 dilution, and serum samples at 1:10 dilution. Fluorescence signals were assessed by indirect immunofluorescence microscopy and graded from negative to positive. Each run included internal positive controls as well as manufacturer-provided positive and negative controls.

### 2.8. Statistical analysis

Non-parametric tests, mainly Mann-Whitney U test and Fishers exact test, are used for analyses in figure 2 comparing demographic and clinical features of autoantibody-positive to negative patients. Correction for multiple comparisons are not used, as no tests are statistically significant ns even without correction for multiple comparisons.

## Results

### 2.9. Study cohort

The study sample consisted of 1021 patients (58% female; median age of 63 (range 34-94)) who were seen at the Karolinska University Hospital memory clinics, Stockholm due to rapid or early-onset memory deterioration at the Karolinska University Hospital memory clinics, Stockholm. The frequency of a confirmed neurodegenerative dementia diagnosis (Alzheimer’s, or Parkinsońs disease-related dementia, fronto-temporal dementia or Lewy-body dementia) in the cohort was 32%, with 51% percent receiving a diagnosis of mild cognitive impairment. The remaining patients most commonly suffered from vascular disorders, affective disorders, fibromyalgia with a few suffering from autoimmune disorders such as multiple sclerosis or systemic lupus erythematosus (Supplementary table 2).

### 2.10. Autoantibody testing

To establish the frequency of neuronal autoantibody seropositivity in the cohort, the patients sera were screened in duplicates for CASPR2, GABAB-R, IgLON5, LGI1 and NMDA-R antibodies by live cell-based assays and scored by microscopy (Table 1). For all antigens apart from NMDA-R, samples that showed a consistent positive pattern in both duplicates were selected for confirmation: 4 positive for CASPR2, 2 for GABAB-R and 2 for LGI1 autoantibodies. As sensitivity for NMDA-R autoantibodies is higher in CSF, even samples that were only positive in one duplicate were also included for further testing. With this lower threshold, a total of 27 suspected NMDA-R positive samples were selected.

**Table 1.** Number of positive samples of all tested samples. All samples were screened with live-CBA tests in serum. Only samples positive at this screening stage were tested further. For NMDA, only samples also positive in CSF were tested further.

| Antigen | CASPR2 | GABAB-R | IgLON5 | LGI1 | NMDA-R |
| --- | --- | --- | --- | --- | --- |
| Live CBA serum | 4 of 1021 | 2 of 1021 | 0 of 1021 | 2 of 1021 | 27 of 1021 |
| Live CBA CSF | 3 of 4 | 2 of 2 | 0 | 0 of 2 | 3 of 25 |
| Fixed CBA serum | 2 of 4 | 0 of 2 | 0 | 1 of 2 | 2 of 3 |
| Fixed CBA CSF | 0 of 2 | 0 of 2 | 0 | 0 of 2 | 1 of 3 |
| TBA serum | 4 of 4 | 2 of 2 | 0 | 1 of 2 | 3 of 3 |
| TBA CSF | 3 of 4 | 0 of 2 | 0 | 1 of 2 | 1 of 3 |
| Confirmed | 3 of 1021 | 2 of 1021 | 0 of 1021 | 1 of 1021 | 2 of 1021 |

For the 4 CASPR2, 2 GABAB-R, 2 LGI1 and 3 NMDA-R patients that were considered potentially positive, live CBA titrations, as well as confirmatory testing with commercial, fixed CBAs as well as rodent brain sections were performed in serum and CSF, (Figure 1, Table 1). A sample was defined as truly positive if confirmed by at least two other tests than the serum live-CBA. For NMDA-R patients, presence of CSF autoantibodies in two independent tests were required, to ensure test specificity, as low-level serum NMDA-R antibodies have been seen in patients with affective disorders, Parkinsońs disease and stroke - all common diagnoses in the cohort (16). This procedure confirmed that eight patients harboured autoantibodies of possible clinical significance against CASPR2 (3), GABAB-R (2), LGI1 (1) and NMDA-R (2; Table 1 and 2). No patients were positive for IgLON5 autoantibodies. Of these patients, three were negative in the commercial autoantibody test (Table 2 and 3).

**Figure 1.**
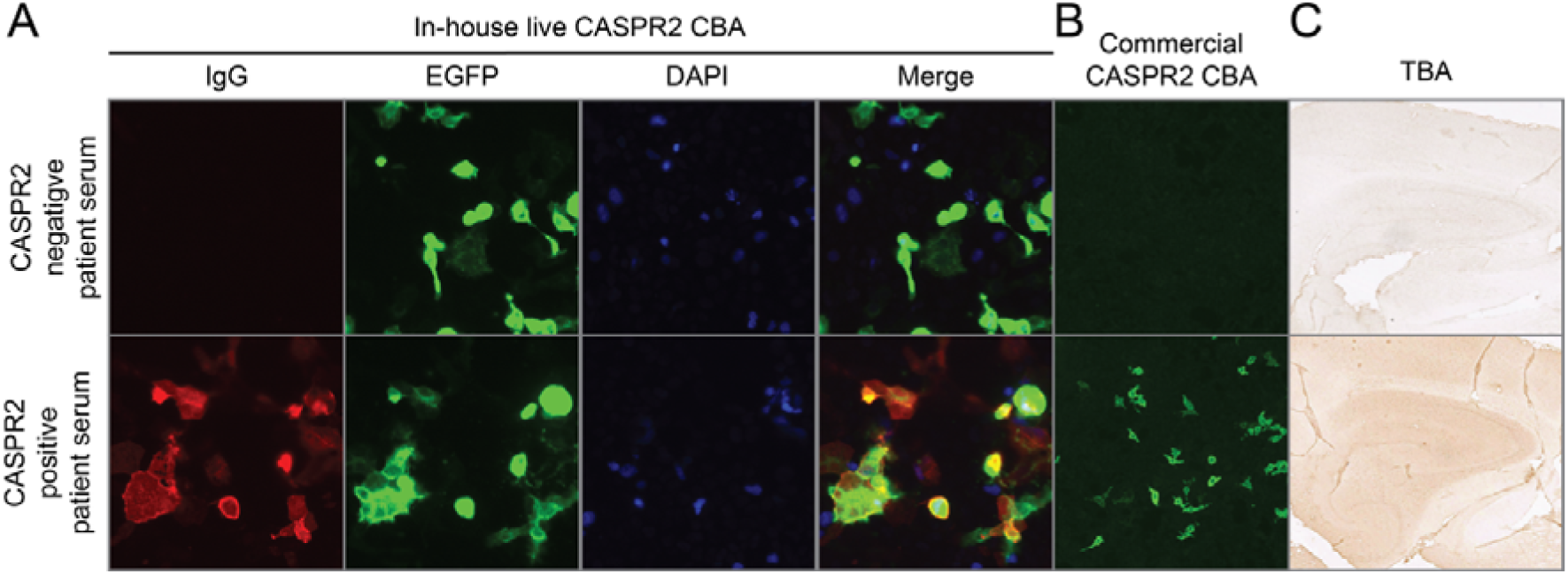
Example of one CASPR-2 negative and one CASPR2-positive serum sample in the in-house live cell-based assay (CBA, A), the commercial CBA (B) and the tissue-based assay (TBA, C), i.e. a horse peroxidase-stained rodent brain section. For the in-house live CBA, green depicts enhanced green fluorescent protein (eGFP) and red depicts IgG staining, whereas for the commercial assay, green depicts IgG staining. DAPI= 4′,6-diamidino-2-phenylindole, a nucleic acid dye depicting cell nuclei.

**Table 2.** Cases with positive test results. EPT = end-point titre, CBA = cell-based assay, TBA = tissue-based assay

| Antigen |  | CASPR2 |  |  | GABAB-R |  | NMDA-R |  | LGI1 |
| --- | --- | --- | --- | --- | --- | --- | --- | --- | --- |
| Case |  | 1 | 2 | 3 | 4 | 5 | 7 | 8 | 6 |
| Live<br>CBA<br>EPT | Serum | 1/3200 | 1/16400 | 1/1600 | >1/320 | 1/160 | 1/320 | 1/160 | 1/80 |
|  | CSF | 1/16 | 1/32 | 1/2 | 1/4 | 1/1 | >1/16 | 1/4 | - |
| Fixed<br>CBA | Serum | + | + | - | - | - | + | + | + |
|  | CSF | NA | NA | - | - | - | + | - | - |
| TBA | Serum | + | + | + | + | + | + | - | + |
|  | CSF | + | + | + | - | - | - | + | + |

**Table 3.** Clinical characterisation of antibody-positive patients. MCI = mild cognitive impairment. MoCA = Montreal cognitive assessment, MMSE = mini mental state examination. *Patient did have seizures at diagnosis <5 years prior. **Neuromyotonia documented at memory clinic but diagnosed later.

| Antigen | CASPR2 |  |  | GABAB-R |  | NMDA-R |  | LGI1 |
| --- | --- | --- | --- | --- | --- | --- | --- | --- |
| Case | 1 | 2 | 3 | 4 | 5 | 7 | 8 | 6 |
| Diagnosis at memory clinic | MCI | MCI | MCI | MCI | MCI | Alzheimer's disease | MCI | LGI1 encephalitis |
| Age at memory clinic visit | >75 | >75 | <55 | <55 | <55 | >75 | >75 | <55 |
| Sex | male | male | male | male | female | female | male | female |
| Cognitive deficits | Yes | Yes | Yes | Yes | Yes | Yes | Yes | Yes |
| MoCA | 28 | NA | 17 | 29 | 27 | 21 | 26 | 25 |
| MMSE | 30 | 27 | 26 | 28 | 30 | 24 | 29 | NA |
| Neurological symptoms | No | Seizures, neuro-pathic pain | Memory gaps | No | No | No | No | No* |
| Neurological examination | Neuro-myotonia** | Normal | Normal | Stiff arms | Normal | Normal | Normal | Normal |
| Malignancy | No | No | No | No | Yes | Yes | Yes | No |
| Comment | - | - | - | - | Cognitive symptoms since breast cancer >5 years prior | Ataxia+Yo+Hu-antibodies leading to breast cancer diagnosis 3 years after | Prostate cancer diagnosed 2 years after | - |

### 2.11. Clinical characterisation of positive cases

Next, the eight antibody-positive cases were further characterised. The patient with LGI1 encephalitis had been previously diagnosed with and treated for AE >5 years prior. As the samples were provided without diagnostic information during the screening, it functioned as an internal, blinded positive control. The remaining seven AE patients had not been investigated for AE prior to or during the memory clinic diagnostic work. Six of these patients received a diagnosis of subjective or mild cognitive impairment, with one NMDA-R antibody-positive patient receiving an Alzheimer’s disorder diagnosis (Table 4). With mild cognitive impairment being the most common diagnosis in the cohort overall, there was no statistically significant difference between the antibody-positive and negative patients given diagnoses (unadjusted Fisher’s test p-value 0.16) (Figure 2A). On a group level, age and sex also did not differ compared to the rest of the cohort (unadjusted Fisher’s test p-values 0.29 and 0.8 respectively) (Figure 2A). The small sample size and heterogeneity of the patient group likely influences this, where for example all three CASPR2 patients were male (Figure 2A), and the two GABABR-encephalitis patients were below 55 years of age, thus more than 15 years below the median of the cohort overall (Figure 2B).

**Figure 2.**
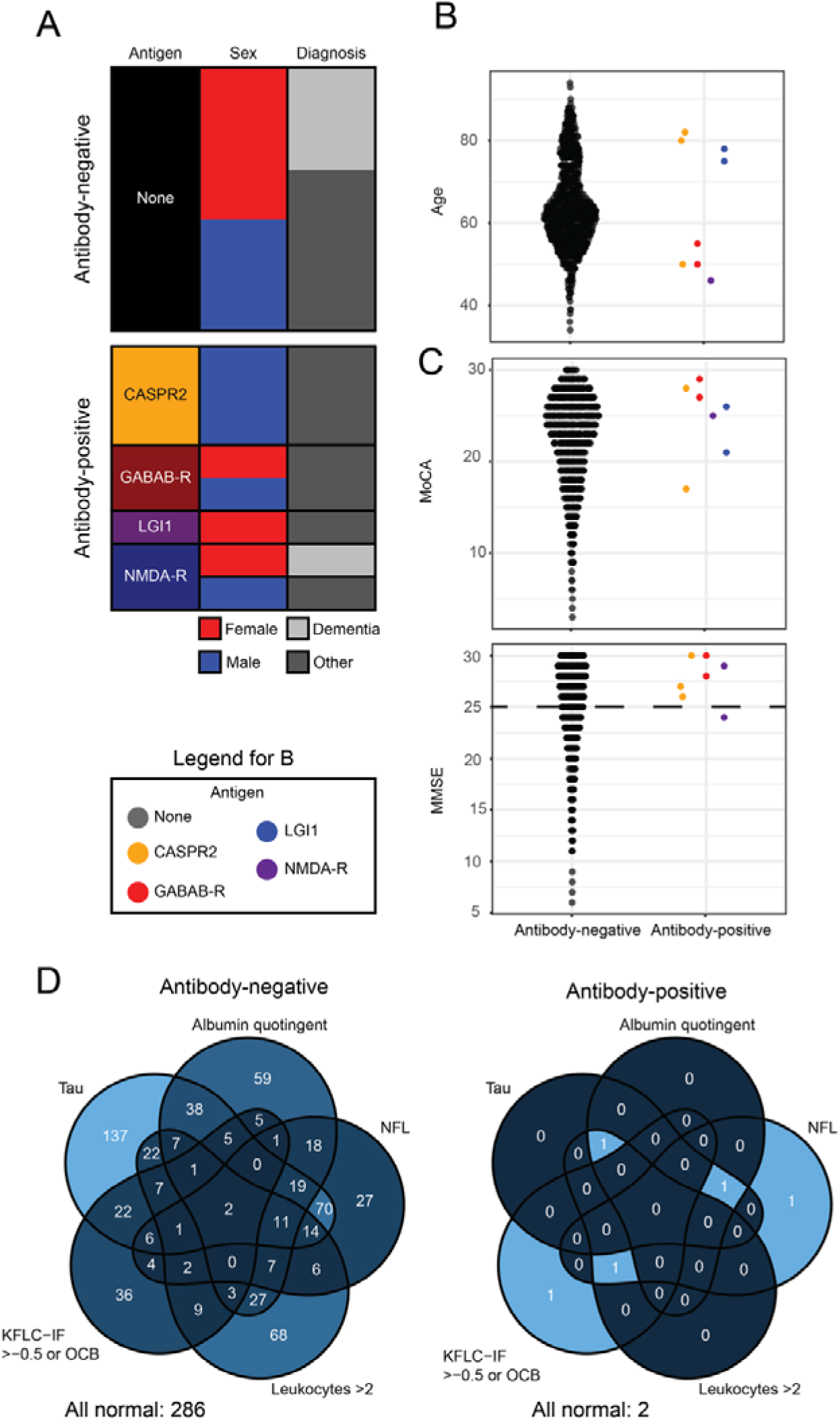
Characteristics of identified patients. A: sex ratio and fraction of dementia diagnoses in the antibody-negative and positive patients. Differences are not significant in either case. B: Age distribution among antibody-negative and positive patients. C: MoCA/MMSE test results assessed close to the time of the lumbar puncture. Differences are not significant in any of the comparisons. Dashed line in MMSE graph indicates threshold for normal tests. For MoCA, the threshold varies depending on age and educational background. D: Venn diagram showing the distribution of the five laboratory tests most likely to differ between autoantibody-negative and positive patients. KFLC-IF = kappa free light chain-intrathecal fraction. OCB: CSF-specific oligoclonal bands in gel electrophoresis. Neither individual laboratory tests, nor the rate of all tests being normal differed significantly between the groups. Due to the low number of autoantibody positive cases and their apparent Venn diagram heterogeneity, no formal statistical analysis of differences in the Venn diagram distribution was attempted.

**Table 4.**
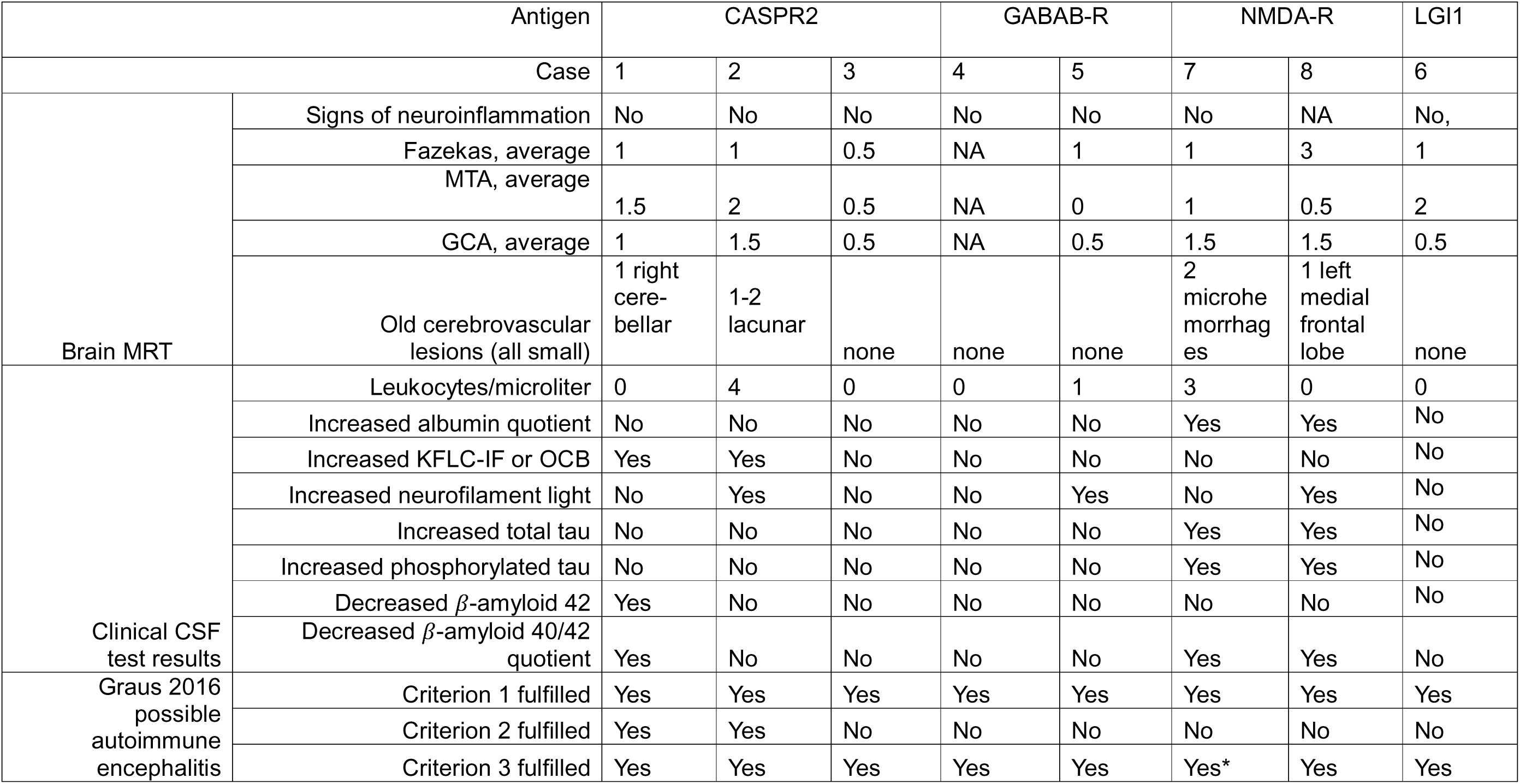
Paraclinical test results, and fulfilment of Graus possible autoimmune encephalitis criteria. KFLC-IF kappa-free light chain-intrathecal fraction, OCB = selectivly observed oligoclonal bands in CSF compared to serum on isoelectric focusing, MRT = magnetic resonance tomography, Fazekas = white mater hyperintensity load score (0-3), indicating grade of small vessel disease. MTA = medial temporal lobe atrophy score (0-4, 0 and 1 normal), GCA = global cortical atrophy scale (0-4). * See dicussion.

| Antigen |  | CASPR2 |  |  | GABAB-R |  | NMDA-R |  | LGI1 |
| --- | --- | --- | --- | --- | --- | --- | --- | --- | --- |
| Case |  | 1 | 2 | 3 | 4 | 5 | 7 | 8 | 6 |
| Brain MRT | Signs of neuroinflammation | No | No | No | No | No | No | NA | No, |
|  | Fazekas, average | 1 | 1 | 0.5 | NA | 1 | 1 | 3 | 1 |
|  | MTA, average | 1.5 | 2 | 0.5 | NA | 0 | 1 | 0.5 | 2 |
|  | GCA, average | 1 | 1.5 | 0.5 | NA | 0.5 | 1.5 | 1.5 | 0.5 |
|  | Old cerebrovascular lesions (all small) | 1 right cerebellar | 1-2 lacunar | none | none | none | 2 microhemorrhages | 1 left medial frontal lobe | none |
| Clinical CSF test results | Leukocytes/microliter | 0 | 4 | 0 | 0 | 1 | 3 | 0 | 0 |
|  | Increased albumin quotient | No | No | No | No | No | Yes | Yes | No |
|  | Increased KFLC-IF or OCB | Yes | Yes | No | No | No | No | No | No |
|  | Increased neurofilament light | No | Yes | No | No | Yes | No | Yes | No |
|  | Increased total tau | No | No | No | No | No | Yes | Yes | No |
|  | Increased phosphorylated tau | No | No | No | No | No | Yes | Yes | No |
| | Decreased β-amyloid 42 | Yes | No | No | No | No | No | No | No |
| | Decreased β-amyloid 40/42 quotient | Yes | No | No | No | No | Yes | Yes | No |
| Graus 2016 possible autoimmune encephalitis | Criterion 1 fulfilled | Yes | Yes | Yes | Yes | Yes | Yes | Yes | Yes |
|  | Criterion 2 fulfilled | Yes | Yes | No | No | No | No | No | No |
|  | Criterion 3 fulfilled | Yes | Yes | Yes | Yes | Yes | Yes* | Yes | Yes |

All seven patients had cognitive complaints. However, only three showed abnormal test results for the Montreal cognitive assessment (MoCA) with one also showing a low score on the mini mental state examination (MMSE). These test results were in line with the overall cohort (unadjusted Mann-Whitney U test p-value 0.19 and 0.1 for MoCA and MMSE, respectively, Figure 2C).

Three showed psychiatric symptoms (affect lability, auditory hallucinations and obsessive compulsions) and two had neurological symptoms (seizures and neuropathic pain and unexplained memory gaps, respectively). Neurological examination showed signs of neuromyotonia in one patient (recorded at the time but diagnosed at examination after CASPR2 antibodies had been found), whereas another showed stiffness in the arms. All other antibody-positive patients had normal examinations.

Two patients had breast cancer (one with GABAB-R antibodies and one with NMDA-R antibodies, later developing anti-Yo and anti-Hu antibodies and ataxia), whereas one NMDA-R-antibody positive patient later developed prostate cancer.

### 2.12. Brain imaging and laboratory test results for positive cases and controls

Next, the paraclinical results for the positive, previously undiagnosed cases were investigated to see if any could be used to find a subset of patients with an increased risk of antibody-positivity.

No patient showed signs of active neuroinflammation on brain magnetic resonance tomography (MRT). One CASPR2 patients showed significant age-adjusted medial temporal lobe atrophy (MTA 2) (13) and three patients had mild to moderate global atrophy (GCA 2). Four patients, all above 75 years of age, showed signs of previous cerebral microhaemorrhages or small infarctions. One of these patients also had a significant white matter lesion load, indicating small-vessel disease (Fazekas 3). Compared to previous results from the cohort, these findings are on average likely less pathological, but with 75% of the overall cohort also show normal results as measured by MTA (13), this absence of degenerative findings is also does not separate the antibody-positive patients from their antibody-negative peers.

We then studied markers relating to neuroinflammation. CSF leukocyte count (a general marker of inflammation), albumin quotient (a sign of blood-brain barrier leakage), kappa-free light chain-intrathecal fraction (KFLC-IF) or CSF-specific oligoclonal electrophoresis bands (OCB, both signs of intrathecal antibody production), neurofilament light (CSF-NFL, axonal damage) or total tau (CSF-Tau, neuronal cell-body damage), were investigated. None of the antibody-positive patients had leukocytes above 4 per microliter and 2-3 patients showed aberrant results for the other parameters of neuroinflammation, with 2/7 showing only normal results. Compared to the rest of the cohort, none of these results were remarkable, either when considered all together or when calculating the likelihood of all being normal (Mann-Whitney U test for the CSF leukocyte count (p=0.33), and Fisher’s exact test for albumin quotient (p=1), KFLC-IF/OCB (p=0.22), CSF-NFL (p=0.37), CSF-Tau (p=0.48) and all 5 tests normal (p = 0.71), no adjustment for multiple comparisons, Table 4).

Next, a wider analysis of all 22 laboratory markers widely available for the cohort was attempted. This included markers of neurodegeneration such as ß-amyloid 42 but also standard blood tests such as Hb, electrolytes and markers of kidney and liver function and revealed no statistical differences (Supplementary table 3).

### 2.13. Fulfilment of Graus 2016 possible autoimmune encephalitis criteria

Finally, it was investigated whether the autoantibody-positive patients fulfilled Graus possible autoimmune encephalitis criteria (9). The first, clinical criterion was considered positive in all cases, given their referral to a centre specialising in memory impairment. Three of the patients showed additional psychiatric and neurological symptoms supporting an encephalitis diagnosis. All patients were assumed to fulfil the alternative diagnoses exclusion criterion, in the case of the patient fulfilling Alzheimer criteria as they showed typical symptoms (ataxia with malignancy and associated paraneoplastic antibodies). However, only two of the CASPR2 antibody-positive patients had paraclinical test results indicative of encephalitis.

## 3. Discussion

In this work, we identify seven (0.7%) undiagnosed individuals with autoantibodies and clinical presentations suggestive of autoimmune encephalitis (AE) among 1021 individuals undergoing dementia diagnostics at the Karolinska University hospital. Five of seven did not fulfil the second, paraclinical Graus criteria for possible autoimmune encephalitis (9). This is not wholly unexpected as multiple studies have reported normal CSF and brain MRI findings in elderly AE patients (7,12,17). In the study of autoimmune encephalitis among patients with suspected neurodegenerative dementia by Bastiaansen et al (7), only 1/7 had had seizures, 2/7 had cell counts above 5 in CSF and none had brain MRI findings typical of ongoing encephalitis. Studies on neurofilament light AE show that 50% of patients with antibodies to LGI1 and as many as 80% of idiopathic or teratoma-associated NMDA-R-encephalitis patients have normal levels of CSF-NFL. With the combination of clinical phenotype and the positive test results in the rigorous autoantibody screening setup, the identified antibody-positive individuals are therefore considered to suffer from autoimmune encephalitis despite this formal lack of criteria fulfilment.

Thus, our study shows that 0.7% of patients at the Karolinska memory clinic have an undiagnosed AE, in line with previous work (7). This finding cannot be generalised to estimate the prevalence of AE in this patient group on a national or international level, for many reasons. Firstly, the Karolinska Memory clinic catchment area for individuals with rapid or early-onset memory impairment is larger than for dementia of any cause, skewing the cohort. Secondly, as many patients undergo brain MRI prior to referral to the memory clinic, individuals with signs of encephalitis might instead be referred to neurology. Thirdly, as our sub-cohort is enriched for individuals with KFLC-IF measurements, it might be skewed to more severe cases; indeed, in work by Rosenberg et al (13) on the same cohort, the rate of dementia was 23%, compared to 33% in our study. Fourthly, it is possible that some cases have been missed with our strategy for defining truly antibody positive cases. Two cases positive for NMDA-R-autoantibodies in serum with the live CBA and the TBA but lacking CSF were e.g. not considered confirmedly antibody-positive, as we prefer to err on the side of caution. With all these caveats in mind, the current AE incidence estimate is merely 3 per million person years in Sweden (8), or 30 new cases per year. If the true prevalence of undiagnosed AE in patients seeking cognitive specialists would be 1/10 of our study result, or 0.07%, with 75 000 unique memory clinic investigations per year (18), these undiagnosed and untreated AE patients would make up more than all conventionally identified cases of AE in the country every year. We believe that this thought experiment underscores the need to identify cost-effective ways to screen for AE in this large population.

Notably, the AE patients did not differ significantly on a group level in any studied variable, clinical or laboratory-based, compared to their autoantibody-negative peers. Even if this is likely due to the small and heterogenous group of AE diagnoses, it still presents a clinical problem; with 2/7 of the undetected AE patients not showing any signs of neuroinflammation or neuronal damage at the time of the lumbar puncture, no available biomarkers have sufficient sensitivity to be used to screen for AE, even without considering specificity. Given that 3/7 of the individuals also did not show clear signs of serum autoantibodies in the commercial fixed cell-based assay, it does imply that a high-sensitivity pre-screening approach for neuroinflammation is needed. Potential markers already in clinical routine in Sweden include CSF levels of IL-6 and CXCL13, that have been shown to be relevant in AE diagnostics (19). Flow cytometry-based B-cell lineage phenotyping could also be potentially useful, given that the CSF compartment of LGI1 and CASPR2 patients have been shown by multiple groups to contain plasmablasts with ultrahigh antigen specificity rates (20)(21). The usefulness of these markers in a memory clinic context would have to be investigated however, as longer time to diagnosis might reduce the active inflammation and therefore the sensitivity of these markers.

In conclusion, autoimmune encephalitis is a rare but clinically significant differential diagnosis to dementia that should be taken into consideration for patients with memory impairment. With insufficient sensitivity of widely used markers of neuroinflammation and issues with sensitivity and specificity of autoantibody screening, identifying feasible biomarkers for AE risk stratification in memory clinics is a goal for coming studies.

## Supporting information

Supplemental tables 1-3 in different tabs

## Data Availability

After the peer-reviewed publication of the manuscript, .nd2 microscopy image files for each sample and antigen will be available for download at Researchdata.se. This database will include information about which samples that have been considered positive in the first screen as well as which samples that were confirmed by other methods, singling out the eight positive cases as "confirmed positive". However, to avoid exposing personal data and enabling full open access to this dataset, it will be fully anonymised and no further information will be available per sample.

## Acknowledgements

The authors are grateful for participation of the patients in the GEDOC cohort. A special aknowledgement is also directed to professor Petter Höglund, Head of Department of Medicine Huddinge, Karolinska Insitutet, for his seminal role in discussions instigating the project.

## Statement of Authorship according to CRediT

**Amanda Freitas Huhtamäki:** Conceptualisation, Methodology, Validation, Investigation, Writing - Original Draft, Writing - Review and Editing.

**Nicole Kleebauer**: Conceptualisation, Methodology, Investigation, Writing - Review and Editing.

**Alexandra Gardner**: Conceptualisation, Methodology, Validation, Writing - Review and Editing.

**Johan Lundberg**: Resources, Writing - Review and Editing.

**Magnus Esbjörnsson**: Writing - Review and Editing.

**Rui Da Silva Rodrigues**: Methodology, Validation, Writing - Review and Editing.

**Patrick Waters**: Methodology, Validation, Writing - Review and Editing.

**Mette Scheller-Nissen**: Methodology, Validation, Writing - Review and Editing.

**Morten Blaabjerg**: Validation, Writing - Review and Editing.

**Nenad Bogdanovic**: Conceptualisation, Resources, Writing - Review and Editing.

**Jakob Theorell**: Conceptualisation, Methodology, Software, Validation, Formal analysis, Data curation, Writing - Original Draft, Writing - Review and Editing, Visualisation, Supervision, Project administration, Funding acquisition.

